# From Delta to Omicron: analysing the SARS-CoV-2 epidemic in France using variant-specific screening tests (September 1 to December 18, 2021)

**DOI:** 10.1101/2021.12.31.21268583

**Authors:** Mircea T. Sofonea, Bénédicte Roquebert, Vincent Foulongne, Laura Verdurme, Sabine Trombert-Paolantoni, Mathilde Roussel, Stéphanie Haim-Boukobza, Samuel Alizon

## Abstract

We analysed 131,478 SARS-CoV-2 variant screening tests performed in France from September 1st to December 18, 2021. Tests consistent with the presence of the Omicron variant exhibit significantly higher cycle threshold Ct values, which could indicate lower amounts of virus genetic material. We estimate that the transmission advantage of the Omicron variant over the Delta variant is +105% (95% confidence interval: 96-114%). Based on these data, we use mechanistic mathematical modelling to explore scenarios for early 2022.

## 1 Introduction

The Omicron SARS-CoV-2 variant of concern (Pango lineage B.1.1.529, Nextstrain clade K21, and GISAID clade GR/484A) was detected in South Africa on November 26, 2021 [1]. Preliminary analyses underline its increased transmission rate [2], high immune evasion potential [3], and low virulence [4,13] compared to the Delta variant.

We analyse Omicron spread in France by applying statistical models to variant-specific screening tests and full genome sequencing. We then use our results to investigate two scenarios regarding the dynamics of French Intensive Care Unit (ICU) occupancy in early 2022.

## 2 Cohort description

The 131,478 screening tests analysed target three mutations in the SARS-CoV-2 Spike protein: E484K (mutation A), E484Q (mutation B), and L452R (mutation C). Therefore, A0B0C1 can correspond to infections caused by the Delta variant, A0B0C0 to the Alpha or Omicron variant, or an ancestral lineage, and A1B0C0 to the Beta or the Gamma variant. The assays used in the study were ID™ SARS-CoV-2/VOC evolution Pentaplex (ID solutions, Grabels, France, 93,554 tests), VariantDetect™ (PerkinElmer, Waltham, USA, 33,037 tests), and VirSNiP (Tib Molbiol, Berlin, Germany, 4,887 tests).

The cohort studied here is described in Table 1. Coverage varies between French regions and the number of tests performed follows the incidence curve of the epidemic (Figure S1).

**Table 1:**
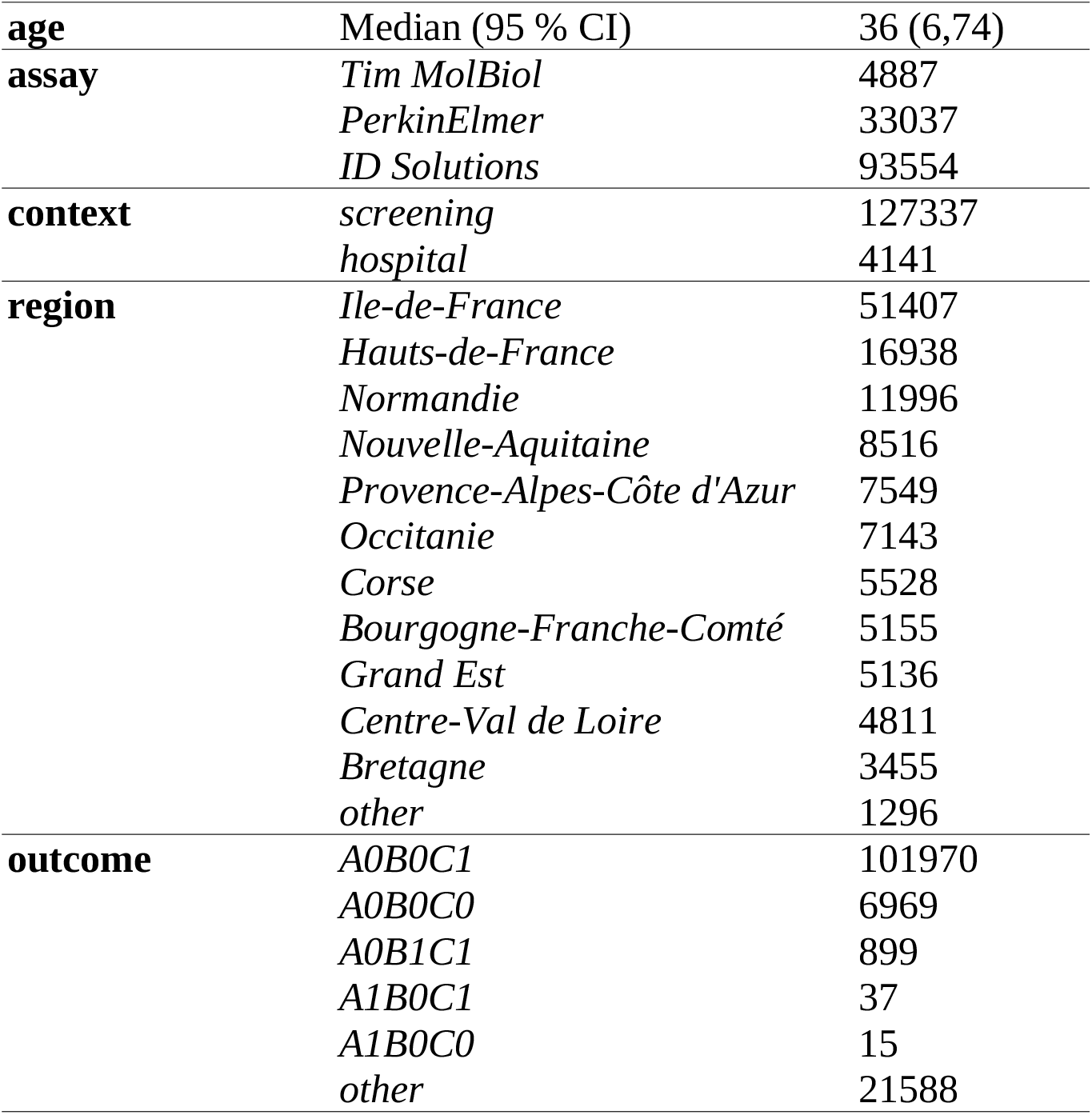
Characteristics of the variant-specific screening tests analysed. (*N* = 131,478). CI stands for confidence interval.

|  |  |  |
| --- | --- | --- |
| **age** | Median (95 % CI) | 36 (6,74) |
| **assay** | *Tim MolBiol* | 4887 |
|  | *PerkinElmer* | 33037 |
|  | *ID Solutions* | 93554 |
| **context** | *screening* | 127337 |
|  | *hospital* | 4141 |
| **region** | *Ile-de-France* | 51407 |
|  | *Hauts-de-France* | 16938 |
|  | *Normandie* | 11996 |
|  | *Nouvelle-Aquitaine* | 8516 |
|  | *Provence-Alpes-Côte d'Azur* | 7549 |
|  | *Occitanie* | 7143 |
|  | *Corse* | 5528 |
|  | *Bourgogne-Franche-Comté* | 5155 |
|  | *Grand Est* | 5136 |
|  | *Centre-Val de Loire* | 4811 |
|  | *Bretagne* | 3455 |
|  | *other* | 1296 |
| **outcome** | *A0B0C1* | 101970 |
|  | *A0B0C0* | 6969 |
|  | *A0B1C1* | 899 |
|  | *A1B0C1* | 37 |
|  | *A1B0C0* | 15 |
|  | *other* | 21588 |

## 3 Factors associated with A0B0C0 tests

Focusing on tests performed between October 25 to December 18, 2021, i.e. when the epidemic was increasing, we used a multinomial regression model to identify factors associated with the result of the variant-specific screening test [5].

A0B0C0 infections were found in younger individuals than A0B0C1 infections (Table 2). We also detected strong temporal increases in most of the French regions with high relative risk ratios (RRR). In some regions, we detected a temporal increase of A0B1C1 tests. Finally, in our dataset, the (rare) A1B0C0 tests were associated with a single region.

**Table 2:** Relative risk ratios (RRR) from the multinomial model. The reference screening result is A0B1C1. 0 indicate non-sognificant values. The model only analyses tests performed after October 25, 2021 (*N* = 103,757). See Table S1 for the tests before that date and Supplementary Methods for additional methodological details.

| **factor** | **value** | **A0B0C0** | **A0B1C1** | **A1B0C0** | **A1B0C1** | **other** |
| --- | --- | --- | --- | --- | --- | --- |
| (Intercept) |  | 0 | 0.01 | 0 | 0 | 0.18 |
| age |  | 0.85 | 1.08 | 0 | 0 | 0.82 |
| context | *screening (ref)* |  |  |  |  |  |
|  | *hospital* | 0 | 0.37 | 0 | 0 | 0.88 |
| assay | *ID Solutions (ref)* |  |  |  |  |  |
|  | *PerkinElmer* | 1.97 | 0.46 | 0 | 0 | 0.82 |
|  | *Tim MolBiol* | 2.06 | 10.85 | 0 | 8.27 | 1.94 |
| date*region | *Ile-de-France* | 86.96 | 4.42 | 0 | 0 | 1.66 |
|  | *BFC* | 10.5 | 8.3 | 0 | 0 | 0.63 |
|  | *Bretagne* | 37.6 | 0 | 0 | 21.54 | 1.33 |
|  | *Centre-Val de Loire* | 46.09 | 0 | 0 | 0 | 0 |
|  | *Corse* | 86.37 | 0.17 | 0 | 0 | 1.92 |
|  | *Grand Est* | 22.18 | 3.67 | 0 | 0 | 0.49 |
|  | *Hauts-de-France* | 44.75 | 0 | 0 | 17.98 | 1.17 |
|  | *Normandie* | 38.23 | 2.17 | 0 | 0 | 0.77 |
|  | *Nouvelle-Aquitaine* | 17.62 | 2.71 | 0 | 0 | 0.43 |
|  | *Occitanie* | 19.83 | 7.7 | 0 | 0 | 0 |
|  | *PACA* | 19.5 | 0 | 0 | 0 | 0.62 |
|  | *other* | 37.59 | 0 | 0 | 0 | 0 |

## 4 Transmission advantages

We then estimated transmission advantages of A0B0C0 infections over A0B0C1 ones during 21 days time windows using methods described in Supplementary Materials and in [6]. The advantage was assumed to be constant over each window. These estimates were corrected for biases in terms of assay used, sampling context (general population vs. hospital), and individual age and administrative region of residency.

In September, A0B0C0 infections were counter-selected compared to A0B0C1 (Figure 1A). This is consistent with the rapid spread of the Delta variant at the time [5]. The pattern changed at the end of November with a 50% transmission advantage, which increased to reach +105% (95% confidence interval: 96.1-114%) in the last time window. According to this model, A0B0C0 infections became more frequent than A0B0C1 ones during the week of December 20 (Figure 1B). Note that the spread of A0B0C0 test results is more advanced in some French regions (Figure S1).

**Figure 1:**
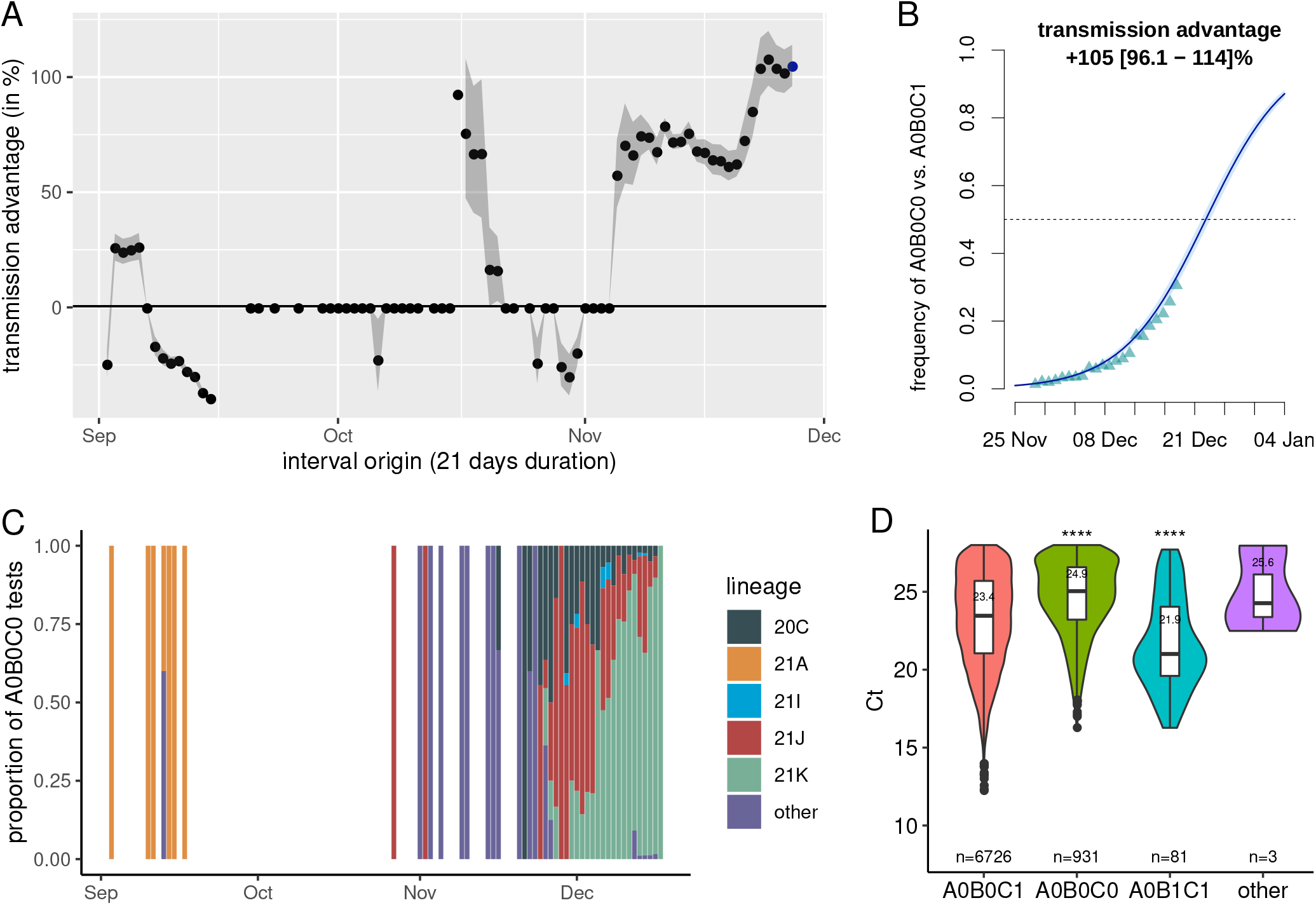
A) Transmission advantage of A0B0C0 tests over A0B0C1 in France, B) Estimated frequency of A0B0C0 relative to A0B0C0 and A0B0C1 tests in France, C) Lineage of the infected associated with A0B0C0 tests using full genome sequencing (*N* = 1,610), and D) Cycle threshold values as a function of the test results (*N* = 7,741). In A, the points indicate the median transmission advantage estimated on a 21 days sliding window and the grey area shows the 95% confidence interval. In B, the triangles show the fitted values from the model, the line the model output, and the shaded area the 95% confidence interval. Panel B corresponds to the last point of panel A. In D, **** indicate a p-value lower than 0.001 in a t.test versus the reference result (i.e. A0B0C1). Only Ct values lower or equal to 28 from tests performed after December 12, 2021 were included.

Using the updated EpiEstim package in R we estimated that, between November 28 and December 18, A0B0C0 infections spread 2.35 (95% CI: 2.28-2.41) times faster than A0B0C1 ones (Figure S2). Since variant screening tests only target 3 mutations, we performed full genome sequencing of 1,160 A0B0C0 samples. Before October 2021, samples were mostly associated with the 21A Nextclade lineage, which is consistent with the Delta variant. In early November, the results were either rare lineages or the 20C lineage. Starting from the end of November, the proportion of A0B0C0 tests linked to the 21K lineage, i.e. the Omicron variant, increased rapidly and, from the second week of December, nearly 75% of the A0B0C0 tests sequenced were caused by Omicron variant at the national level, with, again, spatial heterogeneity across regions (Figure S3).

## 5 Cycle threshold differences

For the *N* = 7,741 screening tests performed with the Perkins kit between December 12 and 18, 2021, we compared the cycle threshold (Ct) values using a linear model with age, administrative region, sampling location, and sampling date as main factors. Only tests with a Ct≤28 were included to minimuse biases in screening test results.

The only factor that was not significant according to an analysis of variance (ANOVA) with a type II error was the sampling location (i.e. general population or hospital). Ct values tended to decrease with age, which is consistent with earlier results [7]. Furthermore, we found that A0B0C0 tests exhibited significantly higher Ct values than A0B0C1 and A0B1C1 tests results, with median values of 24.9 versus 23.4 (Figure 1D). We found a similar trend when using sequencing to assess virus lineage (Figure S4).

This result suggests lower amounts of genetic material in the samples. Note that the high Ct value in the samples labelled as “other” is consistent with these samples yielding non-interpretable results for at least one of the mutations.

## 6 Modelling scenarios

Using the inferred transmission advantage, we explore two scenarios for the beginning of 2022 by updating an earlier epidemiological model tailored to the French epidemic [8], which has been shown to provide robust results on a five weeks horizon [9].

The model follows population natural immunity, which is an output of the model, and vaccine immunity, which is based on national data. The detailed model assumptions are shown in the Supplementary Material. Importantly, we assume that the epidemic reproduction number (*R*_*t*_) grows from 1.08 on December 23, to 1.25 on December 28, and 1.5 on January 7. Furthermore, we also assume that from 15 January, *R*_*t*_ drops to 0.95 due to non-pharmaceutical interventions, and/or spontaneous behavioural changes, and/or contact-network saturation induced by spatial structure [10].

In our ‘optimistic’ scenario, we assume a 3-fold reduction of Omicron virulence compared to Delta, a 75% vaccine efficacy against infection, and 95% against critical forms. In the more ‘pessimistic’ scenario, virulence is only divided by 2 compared to Delta, and vaccine effectiveness is only 40% against infection and 80% against critical forms.

The two scenarios yield a twin-peak pattern(Figure 2). The first peak is identical in both models and corresponds to the peak of the wave of the Delta variant. The second peak is more pronounced in the more pessimistic scenario, where the maximal national ICU capacity of ca. 5,000 beds is exceeded.

**Figure 2:**
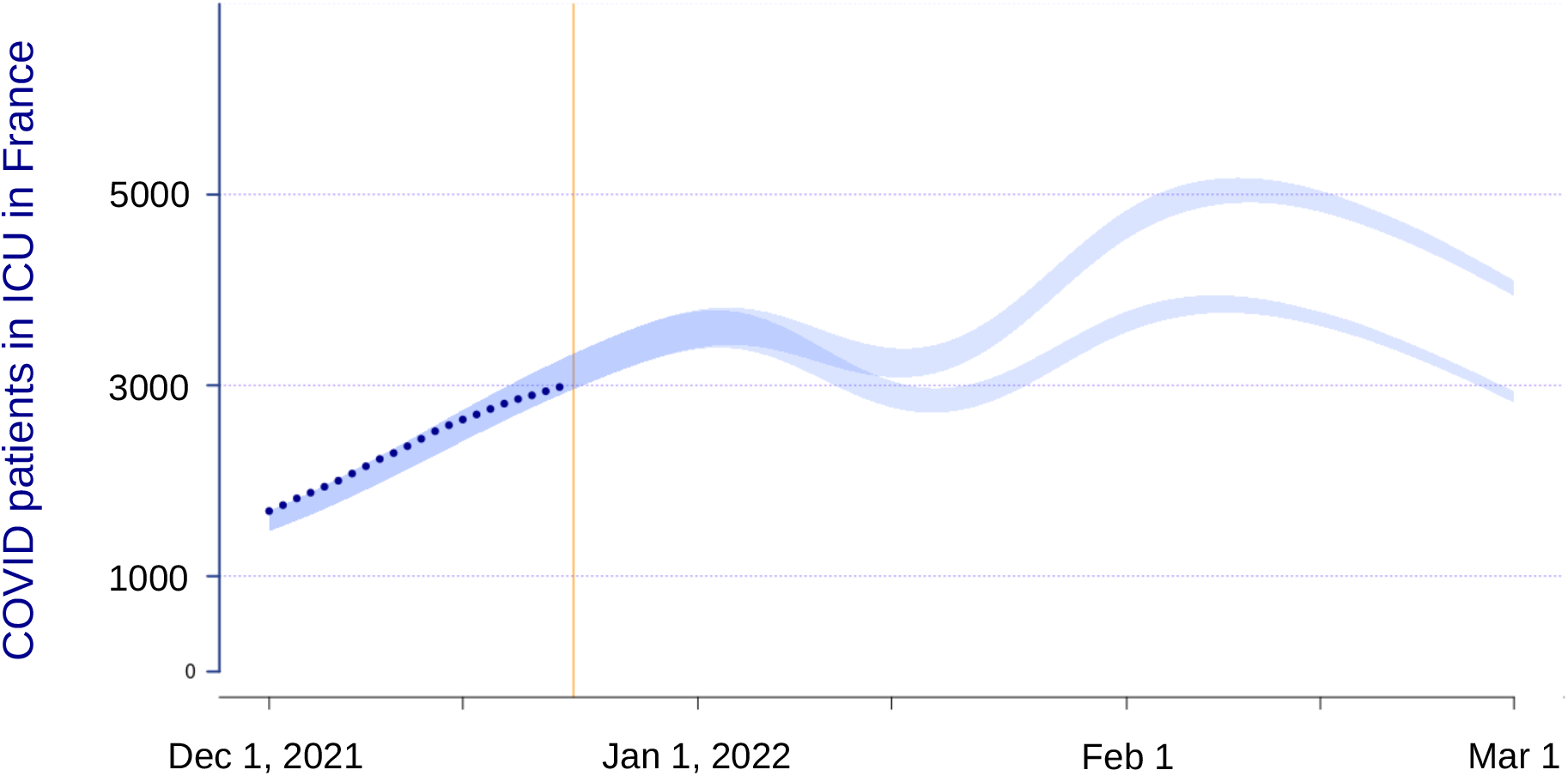
National intensive care unit dynamics in two scenarios of Omicron properties. The vertical yellow line indicates the day the model was performed, the dark blue dot the data, and the shaded envelope the compatibility intervals of the model projections.

## 7 Discussion

Consistently with other countries, we report a rapid spread of the Omicron variant in France. By combining variant-specific screening tests on all positive samples with full-genome sequencing on some of the samples, we show that an increasing proportion of A0B0C0 tests (i.e. without mutations in positions S:484 or S:452) likely corresponds to the spread of the Omicron variant.

A0B0C0 samples exhibited significantly higher cycle threshold (Ct) values than A0B0C1 samples. Although care must be taken when analysing Ct values, especially for coronaviruses [11], this suggests a lower amount of virus genetic material in the samples, which could be consistent with early reports of differences in tissue tropism between the Omicron and Delta variants [12]. This potential lower virus load of the Omicron variant in nasopharyngeal swabs has strong implications regarding RT-PCR test sensitivity.

Finally, epidemiological modelling indicates that even if the virulence of the Omciron variant is reduced compared to that of the Delta variant, the increase in reproduction number we estimate from the data can has the potential to maintain critical COVID-19 activity at a high level in French hospitals, if not overloading them. Therefore, swift mitigation of the epidemic wave appears to be essential.

## Supporting information

Supplementary Methods and Results

## Data Availability

Data will be made available upon publication and are available before that upon reasonable request to the authors.

## Ethical statement

This study has been approved by the Institutional Review Board of the CHU of Montpellier and is registered at ClinicalTrials.gov with identifier NCT04738331.

## Acknowledgements

The authors thank the ETE team from CNRS, IRD, and University of Montpellier for discussion, and the EMERGEN consortium. This project was supported by the Occitanie region and the ANR (PHYEPI grant).

