## Supplementary Methods and Results for "From Delta to Omicron: analysing the SARS-CoV-2 epidemic in France using variant-specific screening tests (September 1 to December 18, 2021)"

December 31, 2021

#### Contents

**S1 Multinomial log-linear model** 2

**S2 Selection advantage estimation** 3

**S3 Cycle threshold analyses** 4

**S4 Mathematical modelling** 5

**Supplementary References** 7

**Supplementary Table** 9

**Supplementary Figures** 10

### S1 Multinomial log-linear model

To perform the multinomial log-linear model, we used the multinom function from the nnet R package. This function uses neural networks to perform model selection in a step wise manner starting from the null model (i.e. without any predictor).

The model formula was the following: `variant   age + assay + location_sampling + date:region`, where `age` is the age of the individual (which is treated as an integer and centred and scaled), `location_sampling` is a binary variable indicating whether the sample was collected in a hospital or not, `assay` corresponds to the type of screening test used, `date` is the sampling date (which is treated as an integer and centred and scaled), and `region` is the French administrative region of sampling.

The multinom function uses an AIC criterion to identify the best model and returns the estimated multinomial logistic regression coefficients as well as their standard error (SE).

These can be used to calculate a $z$-test statistic, which is simply the ratio of the coefficient value to the SE. From there, we can construct a p-value, $P > |z|$, which is the probability the $z$-test statistic would be observed under the null hypothesis and assuming that $z$ follows a normal distribution. Here, we use a classical significance threshold of alpha 5%. When the p-value is smaller than alpha, the null hypothesis can be rejected and the parameter is considered to be significant.

Note that an alternative approach could be to calculate the 95% confidence interval for the coefficient value of the multinomial model using the SE and a critical value on the standard normal distribution.

To give a more intuitive interpretation of the results, we compute the relative risk ratios (RRR) by taking the exponential of the coefficient values of the model. The RRR reflects, for a given variable, how the risk of belonging to one of the outcomes (here variant detection) varies compared to the control group.

Further details about multinomial log-linear models and their interpretations can be found at https://stats.idre.ucla.edu/stata/output/multinomial-logistic-regression/.

#### S2 Selection advantage estimation

Following methods developed in population genetics to estimate the selection coefficient of a mutant allele compared to a wild type allele [1], and following earlier studies in epidemiology [2, 3, 4], we calculate the selection coefficient s by fitting a logistic growth model to the time series of variant frequency. Indeed, provided that the selection coefficient s does not vary over time, and by denoting $p(t)$ the frequency of an allele (here screening test result A0B0C0 or A0B0C1) in the population (here all tests with results A0B0C0 or A0B0C1), we have the following relationship:

$$s = \frac{d}{dt} \log \left( \frac{p(t)}{1 - p(t)} \right) \tag{S1}$$

Note that this value needs to be scaled with respect to the generation time $T$, which is here obtained from the serial interval calculated by **(author?)** [5]. Overall, the transmission advantage $s_T$ of A0B0C0 tests over A0B0C1 tests is given by the formula $s_T = s\,T$.

In order to estimate s, for each region of interest separately, we first perform a generalised linear model (GLM) with a binomial distribution of the residuals (i.e. a logistic regression) where the response variable is the test result (A0B0C0 or A0B0C1) and the factors are the `age` of the individual (which is treated as an integer and centred and scaled), the `assay` used for the test, the sampling `date` (which is treated as an integer and centred and scaled), and the sampling `region`, which is the French administrative region. We then use the fitted values from the GLM to perform the fit of the logistic growth function.

We use 21 days windows to estimate transmission advantage.

We also inferred the transmission advantage using the updated version of the R package EpiEstim [6] available at `https://github.com/mrc-ide/EpiEstim`. We only consider A0B0C0 and A1B1C1 test results and assume that both have the same serial interval (that from [5]).

Further details about the implementation of the inference can be found in the Supplementary R script with the Supplementary data.

#### S3 Cycle threshold analyses

The Perkins variant-specific SARS-CoV-2 tests have a reference probe targeting a region in the N gene of the virus. Following earlier studies [7, 8] use the RT-PCR cycle threshold (Ct) value associated with this reference probe to estimate the amount of genetic material in the sample.

Importantly, care must be taken when interpreting these Ct values. Indeed, the life-cycle of the coronaviruses and the inherent variability of the samples used can generate a large amount of variance [9, 10]. However, here the analyses are performed using the same assay and important covariates such as individual age or sampling region are taken into account.

Statistically, we use a linear model with the following formula:

`Ct    age + variant + location_sampling + date*region` , where `age` is the age of the individual (which is treated as an integer and centred and scaled), `variant` is the outcome of the screening test, `location_sampling` is a binary variable indicating whether the sample was collected in a hospital or not, `date` is the sampling date (which is treated as an integer and centred and scaled), and `region` is the French administrative region of sampling. We also include interactions between the region and the sampling data to capture variations in Ct values that could be linked to differences in epidemic reproduction number. Indeed, growing epidemics can be associated with lower Ct values than declining epidemics [8, 10].

We used a likelihood ratio test to compare this model and a model without the `variant` factor to check that the latter does improve the model.

Factor significance was assessed using an analysis of variance (ANOVA) with a type II error using the Anova function from the car package in R.

The estimated marginal means (EMMs) for the Ct values associated with the screening tests results were computed using the emmeans function from the eponym R package.

### S4 Mathematical modelling

We use the COVIDSIM model, the main structure of which has been described in details in earlier studies [11]. In short, the model is structured in discrete time and uses hospital incidence and prevalence data, as well as mortality data, to estimate key parameters of the COVID-19 epidemic in France. A retrospective analysis showed that the previsions made tend to be robust on a five week window.

In the model, individuals can move between compartments that vary in terms of infectivity. The transition between compartments depends on the time spent in each compartment meaning that the model is non-Markovian (i.e. it captures 'memory' effects).

For simplicity, the transmission model is not stratified by age. However, the probability of developing critical forms, being admitted to critical care, and dying from COVID-19 is age-stratified according to published data [12]. In general, the model parameters correspond to the French epidemic: their initial values are derived from literature data, technical reports or preliminary work, and are then regularly adjusted to reflect hospital dynamics [13].

Vaccination follows the French national campaign data with simplifying assumptions (e.g. immunity is acquired after the second dose). The proportion of people already infected is the result of the model's reconstruction of the epidemic. The immunity of the population can be natural post-infectious (hypothesis of 15% immunologically naïve infected persons, the rest being considered definitively protected), or vaccine-based with, in the most optimistic scenarios, a protection of 80% against infection and 95% against severe forms, without decline over time. Based on our earlier investigations of the French epidemic, we assume that the drop in infectiousness in so-called breakthrough infections in vaccinated individuals is 50%.

The goal of the present model is to explore the impact of the Omicron variant on national intesive care units (ICU) activity. The model is not intended to predict the further but rather to generate trends under assumptions that are arbitrarily optimistic for the most part. In particular, we assume hat

- the set of people vaccinated is homogeneous (no stratification according to the number of doses), which can be interpreted as assuming that the 3rd dose is rapidly generalised to the whole population,

- there is no decrease in vaccine effectiveness over time,
- the proportion of previously infected people that Omicron can reinfect is identical to that of Delta (15%),
- the generation time (number of days between the moment a person is infected and the moment he/she infects another person) is unchanged,
- there are no 'New Year's Eve', 'holiday', 'back-to-school' or meteorological effects on the epidemic spread,
- the epidemic reproduction number (Rt) grows from 1.08 on December 23, 2021, to 1.25 on 28 December, and 1.5 on 7 January, which corresponds to a 10% decrease in the reproduction number at the peak compared to what would be expected by extrapolating the estimated Omicron growth from the screening data,
- from 15 January onwards, a slowdown in the epidemic occurs due to measurements, spontaneous behavioural changes, and/or natural saturation related to spatial structure with a drop in $R_t$ to 0.95
- there is no shortening of the stay in critical care.

We then distinguish two scenarios according to the hypotheses linked to the vaccine immunity and the virulence of the virus:

1. an "optimistic" scenario with a 3-fold reduction in the probability of developing a critical form compared to Delta, a 75% vaccine effectiveness against infection and 95% against critical forms (thus an excellent effect of the 3rd dose),
2. a "pessimistic" scenario where the probability of developing a critical form is only divided by 2 compared to Delta and where the vaccine effectiveness is only 40% against infection and 80% against critical forms.

Again, most of the assumptions made here, e.g. that on January 15, 2022, the epidemic will be decreasing, are rather optimistic. Therefore, the goal of these scenarios is to highlight the potential that, even in such an optimistic configuration, the virulence and reproduction number of the Omicron variant can lead to a saturation of the French ICU capacities.

Table S1 – **Relative risk ratios (RRR) from the multinomial model in the declining early phase of the epidemic ($N = 27,721$).** 0 indicate non-significant values. The reference screening result is A0B1C1. The model only analyses tests performed before October 25, 2021. See Table 1 for the tests after that date.

| factor | value | A0B0C0 | A0B1C1 | A1B0C0 | A1B0C1 | other |
|---|---|---|---|---|---|---|
| (Intercept) | | 0 | 0.01 | 0 | 0.01 | 0.22 |
| age | | 0.78 | 0 | 0 | 0 | 0.94 |
| context | screening (ref) | | | | | |
| | hospital | 0 | 0 | 0 | 0 | 0.74 |
| assay | ID (ref) | | | | | |
| | Tim | 2.94 | 3.31 | 0 | 0 | 1.86 |
| date*region | Ile-de-France | 0.24 | 0 | 0 | 4.68 | 0.73 |
| | BFC | 0.4 | 79145.71 | 2620.66 | 232942.92 | 1.19 |
| | Bretagne | 0.16 | 0 | 1426.58 | 144184.27 | 0.56 |
| | Centre-Val de Loire | 0.15 | 0 | 4424.55 | 0 | 0 |
| | Corse | 0.33 | 198602.42 | 2817.76 | 426501.73 | 1.53 |
| | Grand Est | 0.31 | 0 | 5150.91 | 253786.35 | 1.26 |
| | Hauts-de-France | 0.24 | 0 | 0 | 1641750.51 | 0.87 |
| | Normandie | 0.3 | 2.24 | 0 | 0 | 0 |
| | Nouvelle-Aquitaine | 0.33 | 0 | 0 | 643060.19 | 1.55 |
| | Occitanie | 0.53 | 0 | 2627.52 | 0 | 0.76 |
| | PACA | 0.17 | 3.62 | 0 | 0 | 0.92 |
| | other | 0.27 | 0 | 11633.99 | 451737.39 | 1.58 |

169 # Supplementary Figures

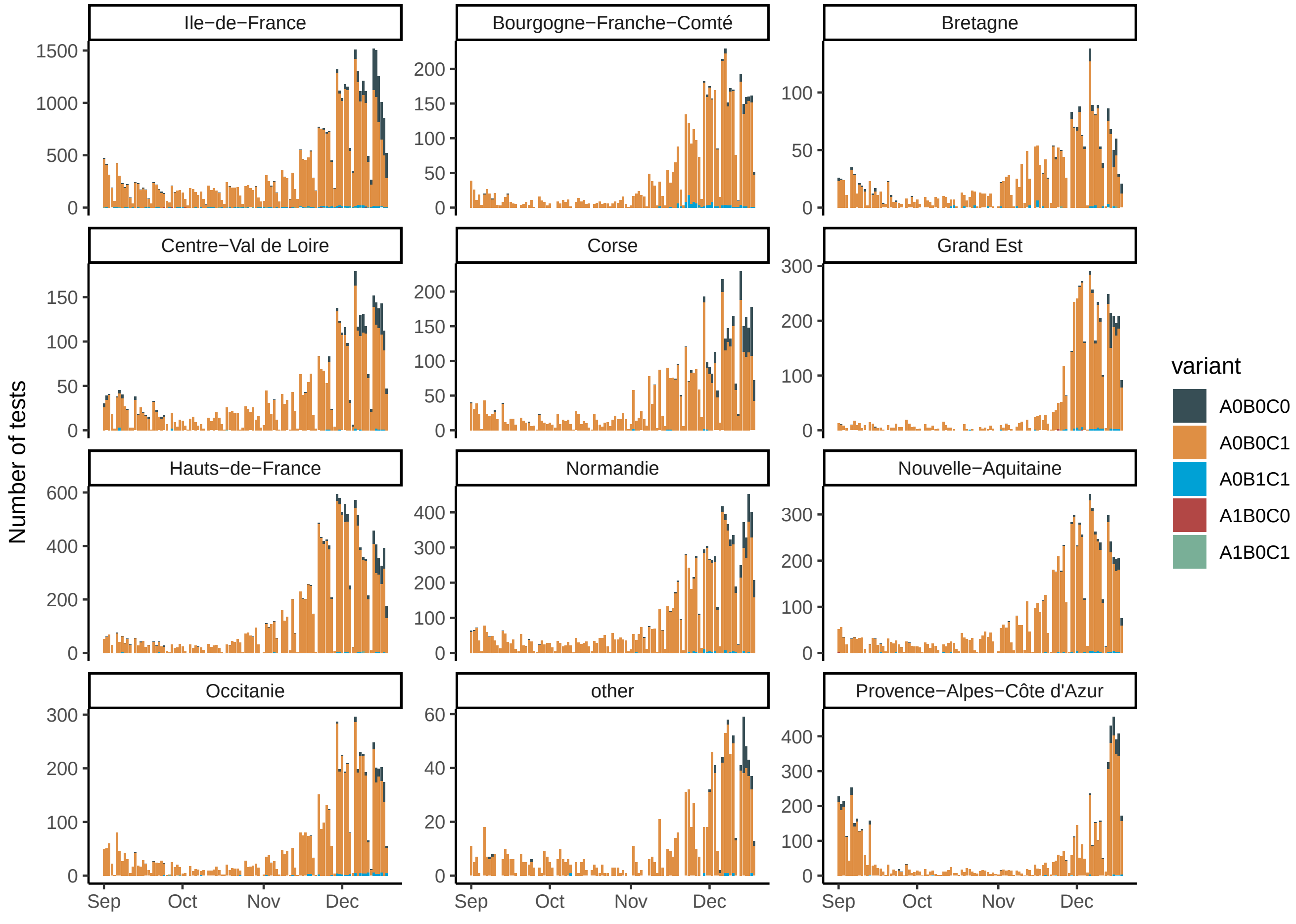


Figure S1 – **Variant-specific screening test incidence data per French region.** The colour indicates the test result. Regions with too few tests are pooled in the "other" category. Notice that the epidemic was declining until mid-October 2021. Furthermore, the proportion of A0B0C0 tests varies across regions.

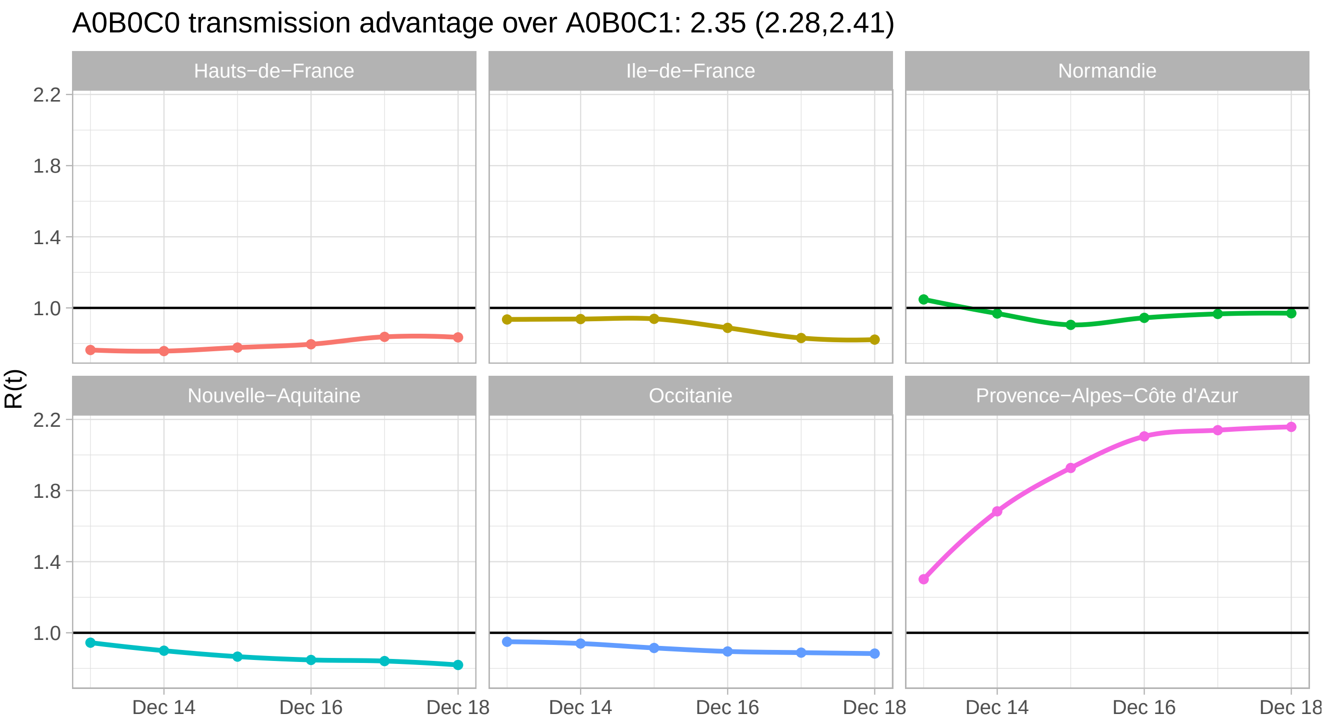


Figure S2 – **Estimation of the temporal reproduction number ($R_t$) and the transmission advantage using the EpiEstim method.** Due to potential delays in test result reporting, since variant-specific screenings adds a delay to the testing, $R_t$ values are likely to be underestimated.

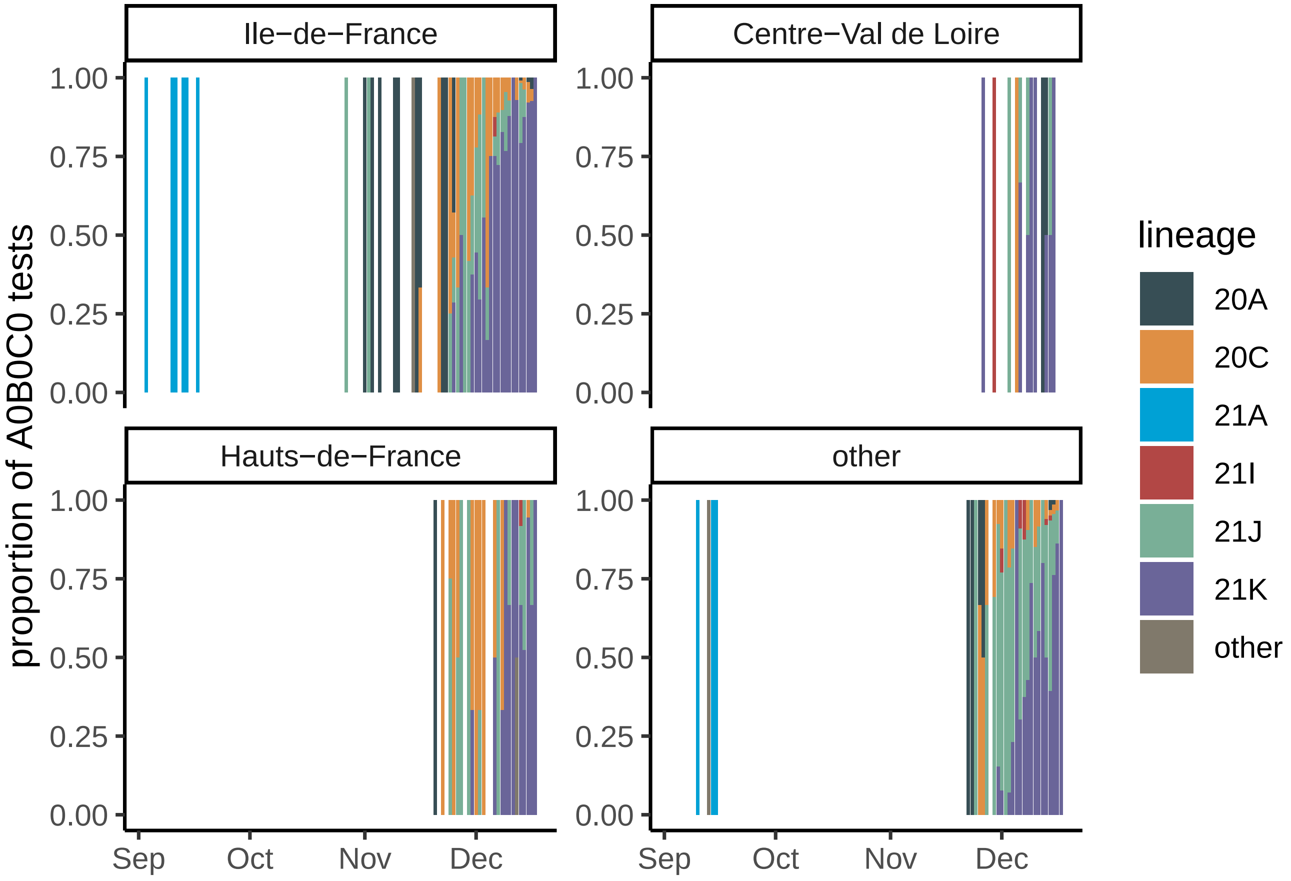


Figure S3 – **Lineage of the infected associated with A0B0C0 tests using full genome sequencing stratified per region ($N = 1,610$).** Regions with less than 50 sequences were pooled in the "other" category. Note that 21K (Omicron) is more prevalent in the Paris area (Ile-de-France region).

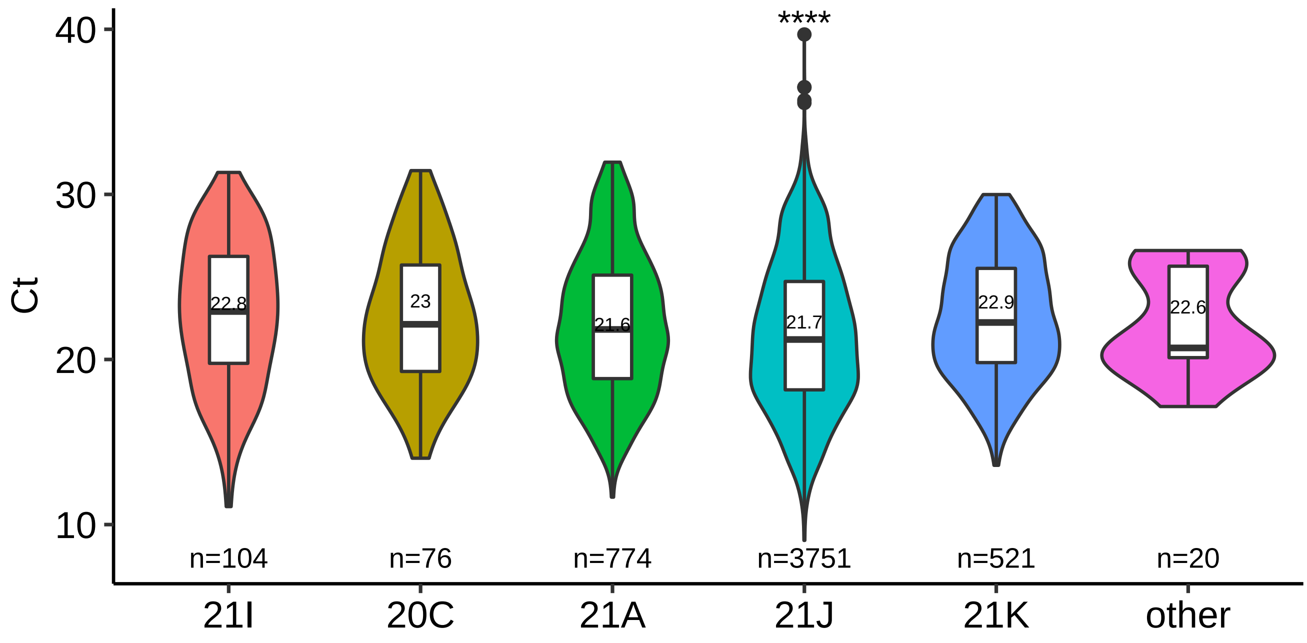


Figure S4 – **Cycle threshold (Ct) value differences as a function of virus Nextclade lineage.** Samples sequences typically have a Ct value lower than 30. The stars indicate a p-value smaller than 0.001 of an t.test using lineage 21K as a reference.
